# Joint effects of sleep duration and depression on the risk of cardiovascular disease and mediating role of depression in middle-aged and older Chinese adults: a nationwide prospective cohort study

**DOI:** 10.1101/2024.10.14.24315497

**Authors:** Shandong Yu, Yanpeng Chu

## Abstract

**Background:** Cardiovascular diseases (CVD) are the leading cause of mortality in China. Depression and sleep disturbances are recognized as risk factors for CVD, but their interplay and combined effects remain unclear. This study examines how sleep duration and depression interact to influence CVD risk.

**Methods:** We analyzed data from five waves of the China Health and Retirement Longitudinal Study (CHARLS) between 2011 and 2020. The final sample included 12,050 individuals aged 45 and above. Sleep duration was categorized as 6 hours or less, 6–8 hours and >8 hours. Depression was assessed using the CESD-10 score and categorized as no depression, mild/moderate depression, and severe depression. Cox proportional hazards models assessed the association between sleep, depression, and CVD. Mediation analysis explored depression’s role in this relationship.

**Results:** Short sleep duration was associated with a higher CVD risk (HR 1.19; 95% CI: 1.03– 1.38). Restricted cubic spline (RCS) analysis showed a U-shaped association between sleep duration and CVD risk, with the lowest risk observed at 7.61 hours of sleep. Severe depression further increased CVD risk when combined with short sleep duration (HR 2.08; 95% CI: 1.46-2.95) or long sleep duration (HR 2.08; 95% CI: 1.05-5.35). Mediation analysis revealed that depression explained 43.8% of the total effect of short sleep on CVD.

**Conclusions:** Both short sleep and depression independently raise the risk of CVD, with depression partially mediating the effect of sleep on cardiovascular health. Interventions targeting both sleep and mental health could reduce CVD risk in high-risk populations.

## Introduction

The prevalence of CVD in China is still on the rise. The mortality of CVD still ranked first, higher than that of tumor and other diseases[1]. Traditional risk factors such as age, cholesterol levels, hypertension, and smoking—components of the Framingham risk score (FRS)—have long been used to predict CVD risk[2, 3]. Despite this, about one-third of individuals with few risk factors still develop CVD, and around 40% of people with low cholesterol levels experience coronary events[2–4]. This underscores the presence of residual CVD risk and highlights the need for additional risk factors to improve risk assessment accuracy[5].

Depression has emerged as a growing global health concern, contributing significantly to the disease burden. The global prevalence of major depressive disorder was estimated at 4.7% in 2013, with an annual incidence rate of 3.0%[6]. It has been reported that up to 23% individuals aged 45 years and older experienced depression symptoms[7]. Several large-scale cohort studies have reported the positive association between depression and CVD[8, 9]. Furthermore, some studies also reported that subclinical depressive symptoms were associated with increased risk of developing cardiovascular disease[10].

Sleep plays a critical role in maintaining physiological homeostasis, and its disturbances have been implicated in the pathogenesis of various cardiovascular conditions, including hypertension, coronary artery disease, and heart failure[11–13]. Short sleep duration and poor sleep quality have been demonstrated associated with increased levels of inflammation, endothelial dysfunction, and sympathetic nervous system activation, all of which are key contributors to CVD[14, 15]. Moreover, sleep disorders such as obstructive sleep apnea have been directly linked to adverse cardiovascular outcomes[16].

The relationship between sleep and depression is bidirectional[17]; individuals with sleep disturbances are more likely to develop depressive symptoms, and those with depression often experience significant sleep impairments[18, 19] This interaction suggests that sleep and depression may not only independently contribute to the development of CVD but also interact synergistically, leading to a higher cardiovascular risk.

Despite the established relationship between sleep, depression, and CVD, studies exploring the combined and interacting impact of sleep and depression on cardiovascular outcomes are limited. This cohort study aims to fill this gap by investigating the combined and interactive effects of sleep and depression on the risk of developing cardiovascular diseases. Understanding these relationships is crucial for developing targeted interventions that address both sleep and mental health, ultimately reducing the burden of cardiovascular disease.

## Methods

### 2.1 Study Population

The data for this study were obtained from the five waves of the China Health and Retirement Longitudinal Study (CHARLS) conducted from 2011 to 2020. CHARLS is a nationally representative longitudinal survey conducted by the National School of Development at Peking University, targeting individuals aged 45 and above, as well as their spouses. The survey assesses the social, economic, and health status of individuals within communities. Participants in CHARLS are drawn from 28 provinces, 150 counties, and 450 communities across China, ensuring national representativeness. CHARLS employs a multi-stage stratified sampling method proportional to size, collecting individual and community characteristics through face-to-face, computer-assisted personal interviews. The study was approved by the Biomedical Ethics Review Committee of Peking University (IRB00001052-11015), and informed consent was obtained from all subjects[20].

Out of the initial 17,708 respondents, to ensure the rigor of the study, we excluded 508 respondents under the age of 45. Additionally, we excluded 243 participants who lacked data on heart problems and stroke, and 2,452 participants who reported heart problems or stroke in 2011. Furthermore, we excluded 1,224 participants who did not answer questions regarding sleep duration and 149 participants who did not participate in the CESD-10 questionnaire. During the follow-up period, 1,082 participants were lost to follow-up. The final sample included in the study was 12,050 participants. We conducted four assessments in 2013, 2015, 2018, and 2020, following the remaining participants for nine years. The detailed inclusion and exclusion process is shown in Figure 1.

**Figure 1.**
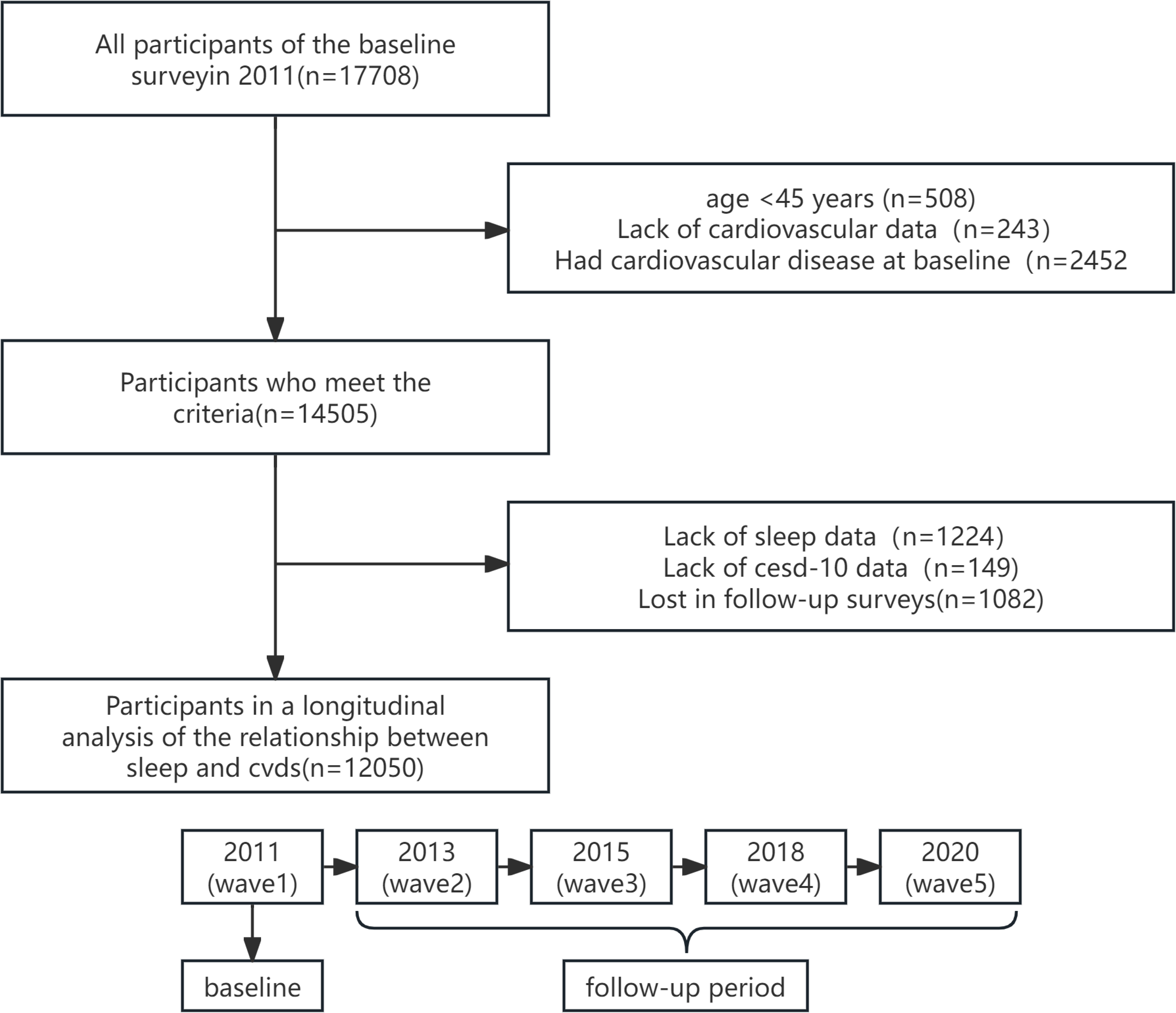
Flowchart and follow-up setting of this study

### 2.2 Measurement of Sleep

Sleep duration was assessed using a self-reported questionnaire: “Over the past month, how many hours of actual sleep did you get at night (on average per night)?” (This may be shorter than the time you spent in bed). Sleep duration was categorized based on the responses, as all reported sleep durations were whole numbers. We defined sleep duration of less than 6 hours as insufficient sleep, 6 to 8 hours as standard sleep, and more than 8 hours as excessive sleep[21].

### 2.3 Measurement of Depression

Depressive symptoms were assessed using the 10-item Center for Epidemiologic Studies Depression Scale (CES-D-10) questionnaire. Individuals with a CES-D-10 score greater than or equal to 20 were classified as having severe depression. Those with a CES-D-10 score less than 20 but greater than or equal to 10 were classified as having mild depression. Individuals with a CES-D-10 score less than 10 were considered normal[22].

### 2.4 Assessment of Cardiovascular and Cerebrovascular Diseases

The assessment of cardiovascular and cerebrovascular diseases was conducted using a self-reported questionnaire: (1) “Have you been diagnosed with a heart attack, coronary heart disease, angina, congestive heart failure, or other heart problems?” and (2) “Have you been diagnosed with a stroke?” The presence of either condition was sufficient to define an individual as having cardiovascular disease[23].

### 2.5 Measurement of Covariates

Sociodemographic characteristics were included as covariates, encompassing age, gender (male and female), body mass index (BMI) (overweight: 24 ≤ BMI < 28, normal: BMI < 24, obese: BMI ≥ 28), educational level (elementary school or below, middle school, high school, college or above), marital status (married or cohabitating, unmarried, separated, divorced, or widowed), and place of residence (rural, urban). Health-related factors previously shown to influence sleep duration and cardiovascular disease, such as smoking status (ever smoked or currently smoking), alcohol consumption (ever drank or currently drinking), hypertension (self-reported hypertension, physical examination showing diastolic blood pressure ≥ 90 mmHg, or systolic blood pressure ≥ 140 mmHg; meeting any criterion was considered hypertension), diabetes or hyperglycemia (self-reported doctor-diagnosed diabetes, blood test showing fasting glucose ≥ 126 mg/dl, or glycated hemoglobin ≥ 6.5%; meeting any criterion was considered diabetes), lung disease (have you ever been told by a doctor that you have a lung disease, excluding tumors or cancer?), kidney disease (have you ever been told by a doctor that you have kidney disease, excluding tumors or cancer?), and dyslipidemia (have you ever been told by a doctor that you have abnormal blood lipids?) were also controlled in this study. These factors were assessed through a self-reported questionnaire.

### 2.6 Statistical Analysis

We stratified descriptive statistics of individual baseline characteristics based on sleep duration. We compared all variables except age using the chi-square test, as they are dichotomous or categorical. We estimated hazard ratios (HR) and their 95% confidence intervals (95% CI) using Cox proportional hazards regression models, with follow-up time as the time scale, to evaluate the association between sleep duration and cardiovascular and cerebrovascular diseases. First, we screened potential confounding factors through univariate regression. Second, we conducted multivariable Cox proportional hazards regression analysis using three models. Model 1 included sociodemographic characteristics such as age and gender. Model 2 extended Model 1 by additionally adjusting for daily living conditions. Model 3 included relevant chronic diseases, namely hypertension, diabetes, lung disease, kidney disease, and dyslipidemia, in addition to the factors in Models 1 and 2. This approach explored the potential impact of these factors on the subjective experiences of different sleep groups. Through this analysis, we aimed to gain a more comprehensive understanding of the relationship between sleep quality and these health variables, thereby providing a basis for future interventions.

Based on prior experience, we hypothesized that depression might mediate the relationship between sleep and cardiovascular disease, so we conducted a mediation analysis to evaluate the direct and indirect associations between sleep, depression, and cardiovascular disease[24]. Sleep (sleep > 8; 8 ≥ sleep > 6; sleep ≤ 6) was used as the predictor variable (X), CES-D-10 score (0-30) as the mediator (M), and the occurrence of cardiovascular disease as the outcome variable (Y). The path model included two parts: the effect of X on M (path a) and the effect of M on Y while controlling for X (path b). The mediation effect, a*b, represents the extent to which the independent variable indirectly influences the dependent variable through the mediator. The significance of the mediation effect was tested using the bootstrap method, with results reported as 95% confidence intervals (CI) and p-values.

Simultaneously, we conducted a joint assessment of cardiovascular risk based on sleep (three categories) and depression (three categories), dividing participants into nine groups. We evaluated the multiplicative and additive interactions of sleep and depression by including an interaction term for sleep and depression in the model to verify the multiplicative effect. Three different indices were used to evaluate the additive interaction: relative excess risk due to interaction (RERI), the proportion of attributable interaction (AP), and the synergy index (SI). RERI = 0, AP = 0, and SI = 1 indicate no interaction, while RERI > 0, AP > 0, and SI > 1 suggest synergistic effects; RERI < 0, AP < 0, and SI < 1 suggest the combined effect is less than the individual effects[25]. The corresponding 95% confidence intervals for these indices were calculated using the delta method[26].

Considering that converting continuous variables into categorical variables is subjective due to the choice of category numbers and cut-off points, which may result in information loss, we also used restricted cubic splines with four knots at the 5th, 35th, 65th, and 95th percentiles to flexibly model the nonlinear association between sleep duration, depression, and cardiovascular and cerebrovascular diseases.

Additionally, we conducted several sensitivity analyses to test the robustness of our findings: 1) Considering the poorer quality of questionnaire responses from individuals with cancer and mental illness, we excluded these individuals. 2) Some participants did not complete all four follow-up rounds; we excluded these individuals to obtain more stable results. 3) Missing covariate data may affect the reliability of the results, so we performed multiple imputations using the “mice” package in R. 4) To assess the dependence of the results on unobserved confounding factors and enhance the credibility of the study conclusions, we used the inverse probability of treatment weighting (IPTW) method to analyze propensity scores.

All statistical analyses were performed using R Studio (version 4.2.1). A two-sided p-value < 0.05 was considered statistically significant.

## Results

### 3.1 Baseline Characteristics of the Population

Using nighttime sleep duration as the grouping variable, we compared baseline differences among the three groups. The results showed statistically significant differences (P < 0.05) between the groups in terms of gender, marital status, place of residence, age, depression, smoking, alcohol consumption, education level, BMI, lung disease, kidney disease, and dyslipidemia, as detailed in Table 1.

**Table 1.**
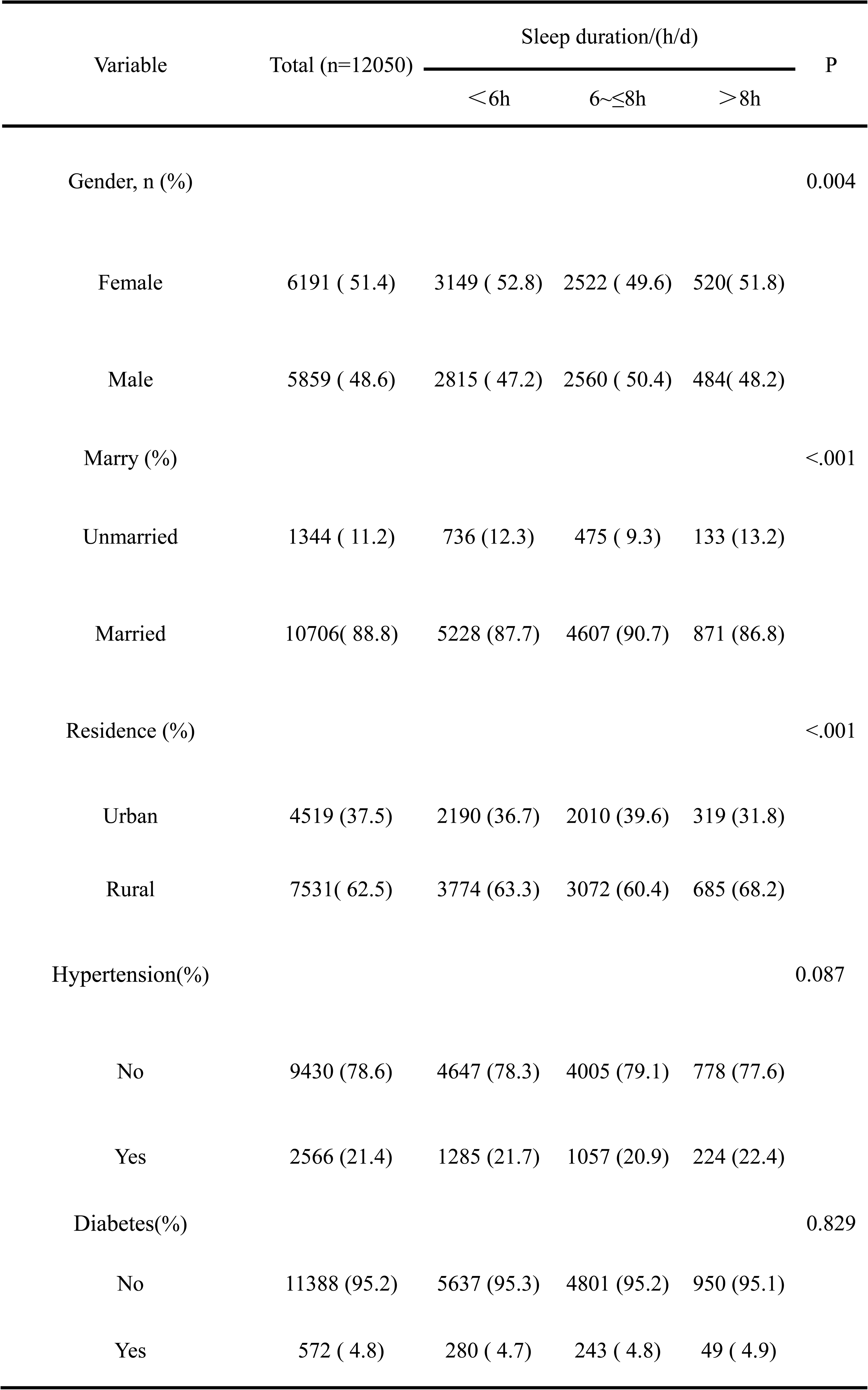
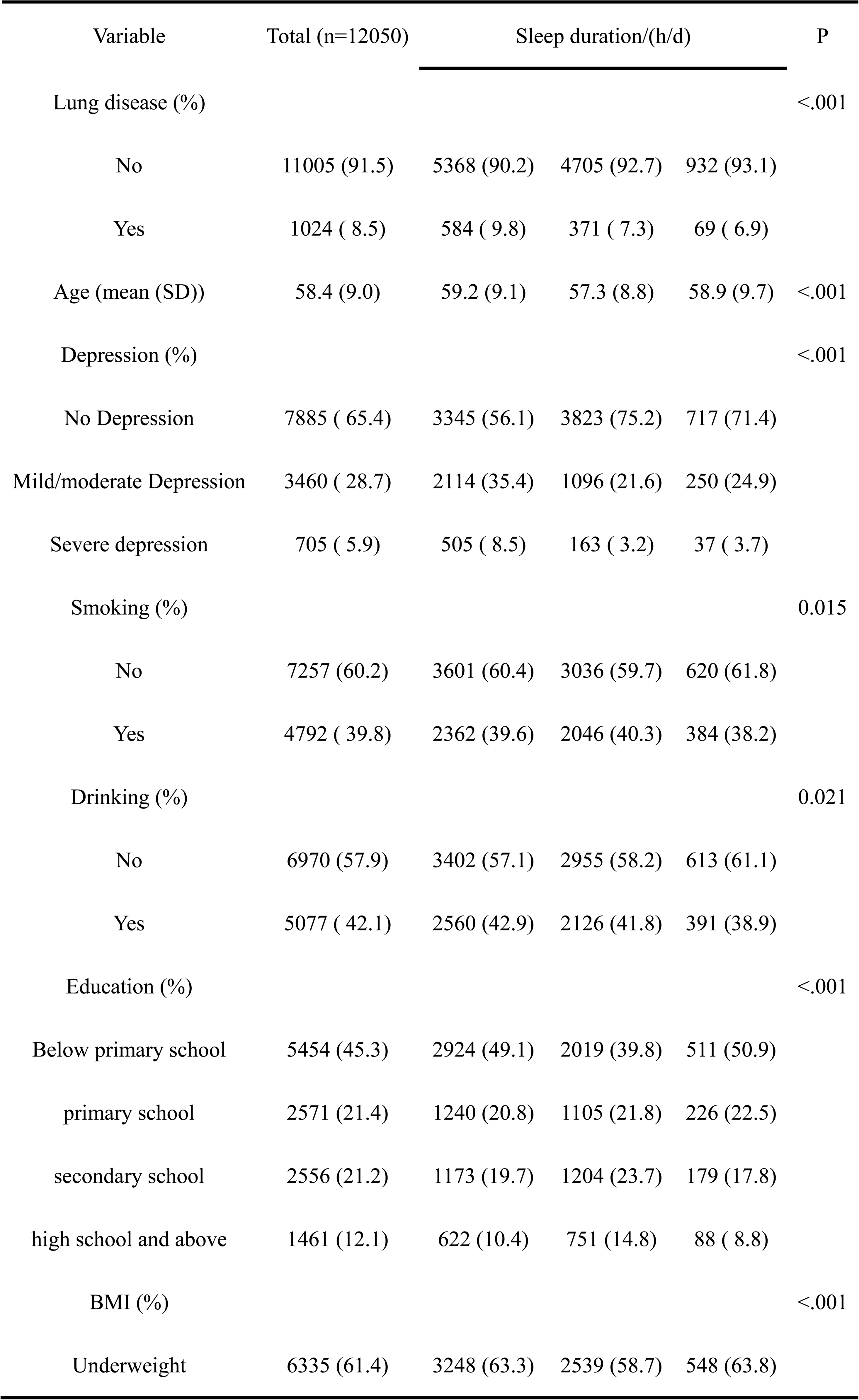
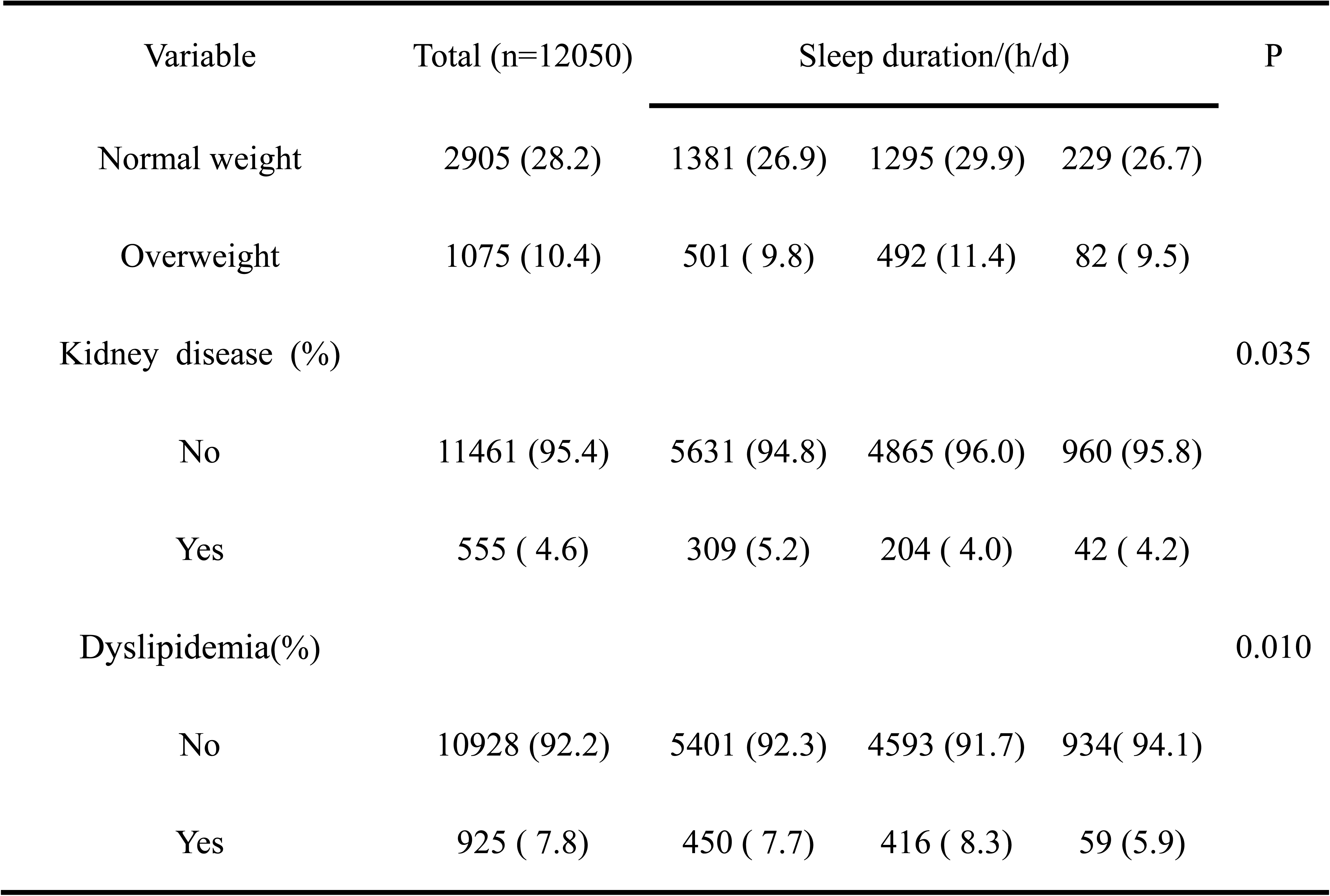
Baseline characteristics of participants according to sleep duration.

| Variable | Total (n=12050) | Sleep duration/(h/d) |  |  | P |
| --- | --- | --- | --- | --- | --- |
|  |  | < 6h | 6~≤8h | > 8h |  |
| Gender, n (%) |  |  |  |  | 0.004 |
| Female | 6191 ( 51.4) | 3149 ( 52.8) | 2522 ( 49.6) | 520( 51.8) |  |
| Male | 5859 ( 48.6) | 2815 ( 47.2) | 2560 ( 50.4) | 484( 48.2) |  |
| Marry (%) |  |  |  |  | <.001 |
| Unmarried | 1344 ( 11.2) | 736 (12.3) | 475 ( 9.3) | 133 (13.2) |  |
| Married | 10706( 88.8) | 5228 (87.7) | 4607 (90.7) | 871 (86.8) |  |
| Residence (%) |  |  |  |  | <.001 |
| Urban | 4519 (37.5) | 2190 (36.7) | 2010 (39.6) | 319 (31.8) |  |
| Rural | 7531( 62.5) | 3774 (63.3) | 3072 (60.4) | 685 (68.2) |  |
| Hypertension(%) |  |  |  |  | 0.087 |
| No | 9430 (78.6) | 4647 (78.3) | 4005 (79.1) | 778 (77.6) |  |
| Yes | 2566 (21.4) | 1285 (21.7) | 1057 (20.9) | 224 (22.4) |  |
| Diabetes(%) |  |  |  |  | 0.829 |
| No | 11388 (95.2) | 5637 (95.3) | 4801 (95.2) | 950 (95.1) |  |
| Yes | 572 ( 4.8) | 280 ( 4.7) | 243 ( 4.8) | 49 ( 4.9) |  |
| Lung disease (%) |  |  |  |  | <.001 |
| No | 11005 (91.5) | 5368 (90.2) | 4705 (92.7) | 932 (93.1) |  |
| Yes | 1024 ( 8.5) | 584 ( 9.8) | 371 ( 7.3) | 69 ( 6.9) |  |
| Age (mean (SD)) | 58.4 (9.0) | 59.2 (9.1) | 57.3 (8.8) | 58.9 (9.7) | <.001 |
| Depression (%) |  |  |  |  | <.001 |
| No Depression | 7885 ( 65.4) | 3345 (56.1) | 3823 (75.2) | 717 (71.4) |  |
| Mild/moderate Depression | 3460 ( 28.7) | 2114 (35.4) | 1096 (21.6) | 250 (24.9) |  |
| Severe depression | 705 ( 5.9) | 505 ( 8.5) | 163 ( 3.2) | 37 ( 3.7) |  |
| Smoking (%) |  |  |  |  | 0.015 |
| No | 7257 (60.2) | 3601 (60.4) | 3036 (59.7) | 620 (61.8) |  |
| Yes | 4792 ( 39.8) | 2362 (39.6) | 2046 (40.3) | 384 (38.2) |  |
| Drinking (%) |  |  |  |  | 0.021 |
| No | 6970 (57.9) | 3402 (57.1) | 2955 (58.2) | 613 (61.1) |  |
| Yes | 5077 ( 42.1) | 2560 (42.9) | 2126 (41.8) | 391 (38.9) |  |
| Education (%) |  |  |  |  | <.001 |
| Below primary school | 5454 (45.3) | 2924 (49.1) | 2019 (39.8) | 511 (50.9) |  |
| primary school | 2571 (21.4) | 1240 (20.8) | 1105 (21.8) | 226 (22.5) |  |
| secondary school | 2556 (21.2) | 1173 (19.7) | 1204 (23.7) | 179 (17.8) |  |
| high school and above | 1461 (12.1) | 622 (10.4) | 751 (14.8) | 88 ( 8.8) |  |
| BMI (%) |  |  |  |  | <.001 |
| Underweight | 6335 (61.4) | 3248 (63.3) | 2539 (58.7) | 548 (63.8) |  |
| Normal weight | 2905 (28.2) | 1381 (26.9) | 1295 (29.9) | 229 (26.7) |  |
| Overweight | 1075 (10.4) | 501 ( 9.8) | 492 (11.4) | 82 ( 9.5) |  |
| Kidney disease (%) |  |  |  |  | 0.035 |
| No | 11461 (95.4) | 5631 (94.8) | 4865 (96.0) | 960 (95.8) |  |
| Yes | 555 ( 4.6) | 309 (5.2) | 204 ( 4.0) | 42 ( 4.2) |  |
| Dyslipidemia(%) |  |  |  |  | 0.010 |
| No | 10928 (92.2) | 5401 (92.3) | 4593 (91.7) | 934( 94.1) |  |
| Yes | 925 ( 7.8) | 450 ( 7.7) | 416 ( 8.3) | 59 (5.9) |  |

### 3.2 Identification of Potential Confounders

Using the occurrence of cardiovascular events as the outcome, we constructed univariate Cox regression models. The results indicated that gender, age, marital status, place of residence, hypertension, diabetes, alcohol consumption, education level, BMI, depression, lung disease, kidney disease, and dyslipidemia were associated with the occurrence of the outcome (P < 0.05), as detailed in Table 2.

**Table 2.** Univariate cox model.

| Variables | $\beta$ | S. E | Z | P | HR (95%CI) |
| --- | --- | --- | --- | --- | --- |
| Gender |  |  |  |  |  |
| Female |  |  |  |  | Ref |
| Male | -0.173 | 0.030 | -5.816 | <.001 | 0.841 (0.794, 0.892) |
| Age | 0.015 | 0.002 | 8.464 | <.001 | 1.015 (1.012, 1.019) |
| Marry |  |  |  |  |  |
| Non-married |  |  |  |  | Ref |
| Married | -0.291 | 0.037 | -7.899 | <.001 | 0.748 (0.696, 0.804) |
| Residence |  |  |  |  |  |
| Urban |  |  |  |  | Ref |
| Rural | -0.111 | 0.030 | -3.670 | <.001 | 0.895 (0.843, 0.950) |
| Hypertension |  |  |  |  |  |
| No |  |  |  |  | Ref |
| Yes | 0.821 | 0.030 | 27.410 | <.001 | 2.272 (2.142, 2.409) |
| Diabetes |  |  |  |  |  |
| No |  |  |  |  | Ref |
| Yes | 0.622 | 0.036 | 17.230 | <.001 | 1.862 (1.735, 1.999) |
| Smoking |  |  |  |  |  |
| No |  |  |  |  | Ref |
| Yes | -0.006 | 0.041 | -0.141 | 0.888 | 0.994 (0.916 - 1.079) |
| Drinking |  |  |  |  |  |
| No |  |  |  |  | Ref |
| Yes | -0.493 | 0.037 | -13.260 | <.001 | 0.611 (0.568, 0.657) |
| Lung disease |  |  |  |  |  |
| No |  |  |  |  | Ref |
| Yes | 0.652 | 0.034 | 18.960 | <.001 | 1.920 (1.795, 2.054) |
| Education |  |  |  |  |  |
| Below Primary school |  |  |  |  | Ref |
| Primary school | 0.010 | 0.038 | 0.266 | 0.791 | 1.010 (0.937-1.089) |
| Secondary school | -0.057 | 0.039 | -1.456 | 0.145 | 0.944 (0.8745-1.020) |
| High school and above | 0.070 | 0.047 | 1.489 | 0.136 | 1.072 (0.978-1.175) |
| BMI |  |  |  |  |  |
| Underweight |  |  |  |  | Ref |
| Normal weight | 0.321 | 0.071 | 4.534 | <.001 | 1.379 (1.200 -1.584) |
| Overweight | 0.630 | 0.089 | 7.103 | <.001 | 1.878(1.578 - 2.235) |
| Depression |  |  |  |  |  |
| No depression |  |  |  |  | Ref |
| Mild/moderate depression | 0.251 | 0.033 | 7.630 | <.001 | 1.285 (1.205 - 1.371) |
| Depression | 0.652 | 0.047 | 13.830 | <.001 | 1.920 (1.750 - 2.106) |
| Kidney disease |  |  |  |  |  |
| No |  |  |  |  | Ref |
| Yes | 0.940 | 0.040 | 23.690 | <.001 | 2.559 (2.368-2.766) |
| Dyslipidemia |  |  |  |  |  |
| No |  |  |  |  | Ref |
| Yes | 1.266 | 0.031 | 41.530 | <.001 | 3.548 (3.342-3.767) |

### 3.3 Association Analysis of sleep duration and cardiovascular disease

We constructed a Cox regression model with sleep duration as the independent variable and the occurrence of cardiovascular events as the dependent variable. Model 1 adjusted for two demographic variables: gender and age. Model 2 further adjusted for daily living conditions, while Model 3 additionally adjusted for disease data. Differences between the ≤6 hours group and the 6 to ≤8 hours group were statistically significant across all three models, with p-values of <0.001, 0.008, and 0.019 for Model 1, Model 2, and Model 3, respectively. Differences between the >8 hours group and the 6 to ≤8 hours group were not statistically significant in Model 1, Model 2, or Model 3 (P > 0.05). According to Model 3, compared to individuals who slept 6 to 8 hours per night, those who slept less than 6 hours showed a higher risk of event occurrence, with a hazard ratio (HR) of 1.19 (95%CI:1.03-1.38), indicating a 19% increase in risk. In contrast, individuals who slept more than 8 hours did not show a significant difference in risk compared to the reference group, with an HR of 0.97 (95%CI: 0.74-1.26). Overall, the results from Model 3 indicate that insufficient sleep is significantly associated with a higher risk, while long sleep duration does not have a significant impact on the risk of event occurrence (Figure 2).

**Figure 2.**
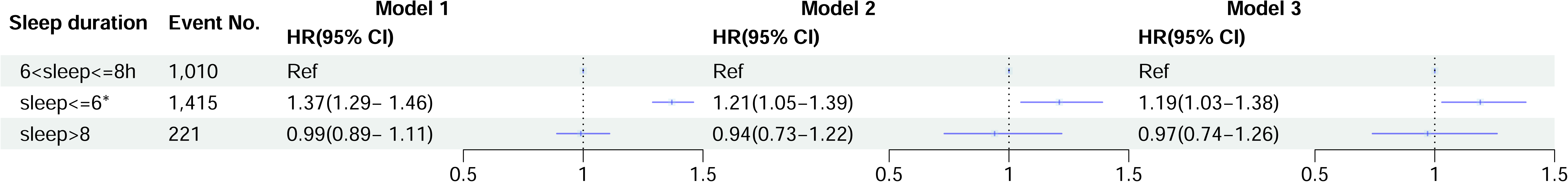
Multi-model Cox regression. Model 1: adjust age, gender; Model 2: adjust age, gender, marital status, residence, smoking, drinking, BMI, depression, education.; Model 3: adjust as Model 2 plus dyslipidemia, kidney disease, diabetes, hypertension, lung disease. The number of events for each group is also provided. Asterisks (*) denote significant associations with P-values < 0.05.

### 3.4 RCS Plot and Histogram

After adjusting for potential confounders, Restricted Cubic Spline (RCS) regression revealed a nonlinear association between sleep duration and cardiovascular events (P < 0.001). The results indicated that sleep durations of less than 6 hours were associated with an increased risk of cardiovascular disease (CVD). In contrast, sleep durations between 6 and 10 hours were associated with a reduced risk of CVD, showing a U-shaped distribution. The lowest risk of CVD was observed at a sleep duration of 7.61 hours. However, when sleep duration exceeded 10 hours, the risk of CVD increased with longer sleep durations. Depression showed a linear and positive correlation with cardiovascular events, although it was not statistically significant (P = 0.629) (Figure 3).

**Figure 3.**
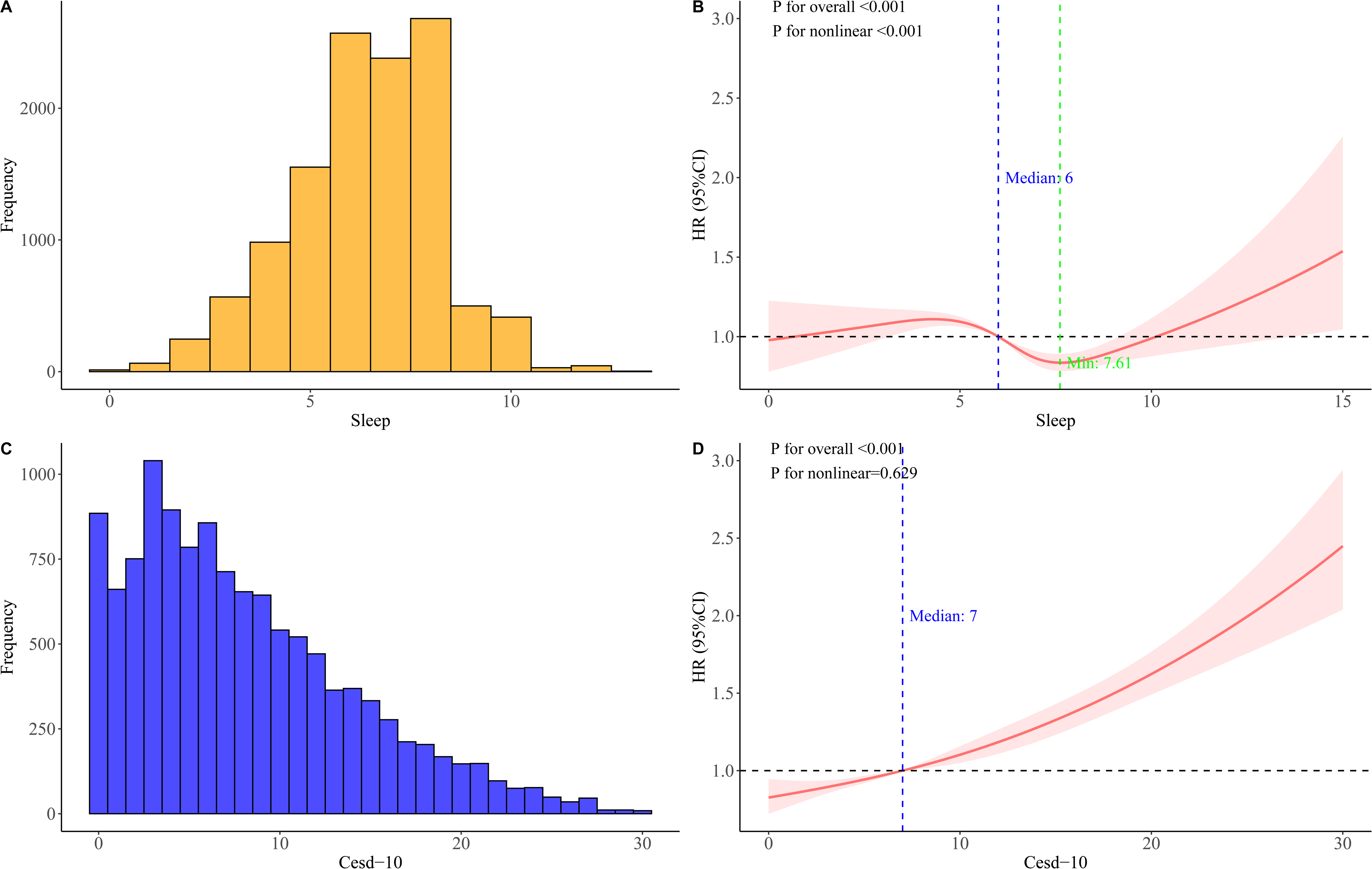
Associations of sleep and depression with cardiovascular disease (CVD). A, C show the distribution for sleep duration and CESD-10 score; B, D present RCS curves showing hazard ratios (HRs) for CVD, adjusted for age, gender, marital status, residence, smoking status, drinking status, body mass index (BMI), depression, education level, dyslipidemia, kidney disease, diabetes, hypertension, and lung disease. Data were fitted using Cox proportional hazards regression models. Solid lines represent HRs, and shaded areas indicate 95% confidence intervals (CIs). CI, confidence interval; CESD-10, Center for Epidemiologic Studies Depression Scale-10; RCS, restricted cubic spline.

### 3.5 Mediation Analysis of Depression

We also examined the mediating role of depression in the pathway between sleep duration and cardiovascular disease (CVD). The analysis revealed a total effect of −0.011, The indirect effect was −0.005, The direct effect was −0.006. The results indicated that depression played a crucial mediating role in this pathway, contributing to 43.8% of the total effect. This finding suggests that sleep duration reduces CVD risk both directly and by alleviating depression, highlighting the complex relationship between sleep, depression, and cardiovascular disease. These results suggest that improving sleep may not only offer direct physiological benefits but also reduce the risk of cardiovascular disease by mitigating depression (Table 3, Supplemental Figure 1).

**Table 3.** Mediation analysis.

| Type | Estimate | 95% CI | P |
| --- | --- | --- | --- |
| Total Effect | -0.011 | -0.014,-0.009 | <0.001 |
| ACME | -0.005 | -0.006,-0.004 | <0.001 |
| ADE | -0.006 | -0.009,-0.004 | <0.001 |
| Proportion Mediated | 0.438 | 0.359,0.550 | <0.001 |

### 3.6 Interaction and combined effects of sleep and depression on cardiovascular disease incidence

No significant multiplicative and additive interactions were found between sleep duration and depression on cardiovascular disease incidence (Additive: RERI = 0.04, 95% CI −0.04–0.12; AP=0.04, 95% CI −1.90–1.97; SI= 2.29, 95% CI −0.22–4.80; Multiplicative, HR = 1.07, 95% CI 0.90–1.27) (Table 4). Figure 4 shows the joint association of sleep duration and depression on the primary outcomes. HRs for individuals of sleep duration <6h and severe depression compared with those with sleep duration between 6-8h and no depression was 1.71 (95% CI: 1.26–2.32) for CVD incidence after adjusting for confounders. In addition, among different sleep duration subgroups, CVD risk is increased by depression.

**Figure 4.**
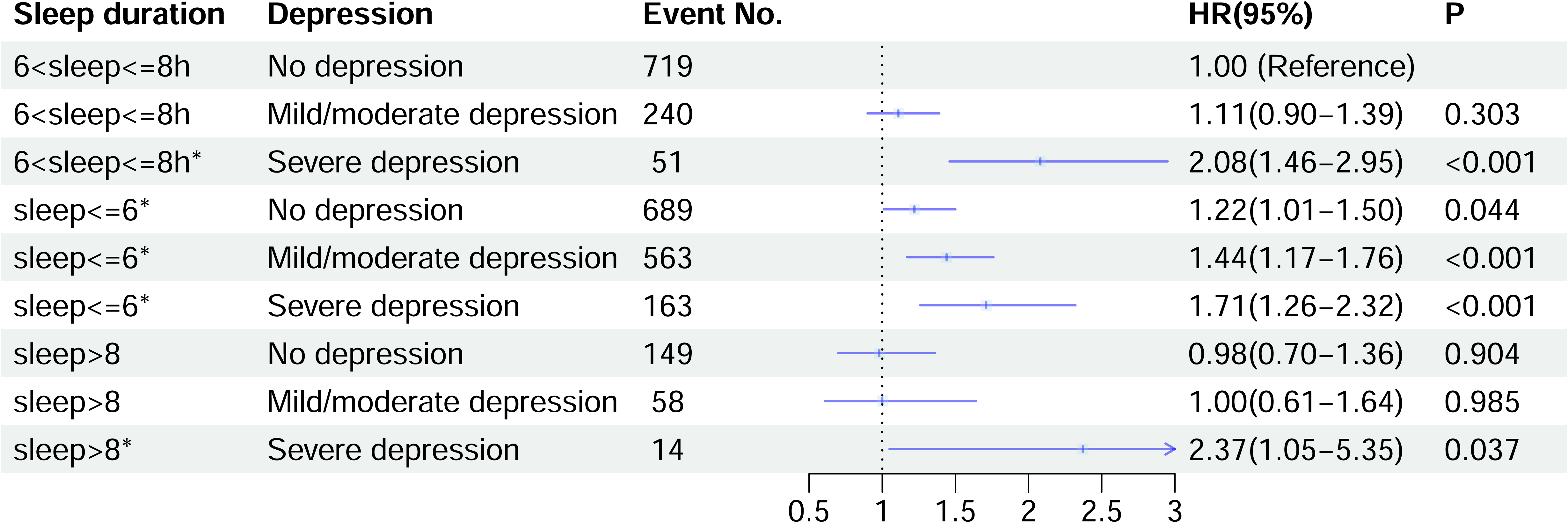
Joint effects of sleep duration and depression on CVD risk. Sleep duration is categorized into three groups: ≤6 hours, 6 to ≤8 hours (reference group), and >8 hours. Depression is categorized into three levels: no depression, mild/moderate depression, and severe depression. The plot illustrates how the combination of short or long sleep duration with varying levels of depression impacts cardiovascular risk. Significant associations are observed particularly in those with severe depression across different sleep duration categories. The number of events for each group is also provided. Asterisks (*) denote significant associations with P-values < 0.05.

**Table 4.** Interactive effects of sleep duration and depression.

| Interactive items | Interactive effects (95% CI) |
| --- | --- |
| Additive effects |  |
| RERI | 0.04 (-0.04 - 0.12) |
| AP | 0.04 (-1.90 - 1.97) |
| SI | 2.29(-0.22 - 4.80) |
| Multiplicative effect | 1.07 (0.90 - 1.27) |

### 3.7 Subgroup and sensitivity analysis

To investigate the effects of sleep duration on various populations, we conducted a subgroup analysis based on age, gender, hypertension, and diabetes status. Our findings indicate that short sleep duration significantly increases the risk of cardiovascular disease (CVD) across all subgroups(Table 5).

**Table 5.** Subgroup analysis.

| Variables | $\beta$ | S. E | Z | P | HR (95%CI) |
| --- | --- | --- | --- | --- | --- |
| Gender |  |  |  |  |  |
| Gender=Male |  |  |  |  |  |
| 6~≤8h |  |  |  |  | Ref |
| ≤6 | 0.18 | 0.05 | 3.31 | <.001 | 1.26 (1.01- 1.57) |
| >8h | 0.13 | 0.09 | 1.50 | 0.134 | 0.90 (0.62, 1.30) |
| Gender=Female |  |  |  |  |  |
| 6~≤8h |  |  |  |  | Ref |
| ≤6 | 0.14 | 0.05 | 1.46 | <.001 | 1.20 (1.09, 1.33) |
| >8h | -0.43 | 0.09 | -0.60 | 0.551 | 0.95 (0.79, 1.14) |
| Age |  |  |  |  |  |
| Age<65 |  |  |  |  |  |
| 6~≤8h |  |  |  |  | Ref |
| ≤6 | 0.20 | 0.05 | 3.80 | <.001 | 1.22 (1.10-1.36) |
| >8h | 0.17 | 0.10 | 1.70 | 0.089 | 1.18 (0.98-1.43) |
| Age≥65 |  |  |  |  |  |
| 6~≤8h |  |  |  |  | Ref |
| ≤6 | 0.16 | 0.05 | 3.17 | <.001 | 1.18(1.06-1.30) |
| >8h | -0.06 | 0.08 | -0.66 | 0.510 | 0.95(0.80-1.12) |
| Hypertension |  |  |  |  |  |
| Hypertension=NO |  |  |  |  |  |
| 6~≤8h |  |  |  |  | Ref |
| ≤6 | 0.29 | 0.05 | 5.79 | <.001 | 1.33(1.21-1.47) |
| >8h | 0.21 | 0.09 | 0.23 | 0.82 | 1.02(0.85-1.22) |
| Hypertension=Yes |  |  |  |  |  |
| 6~≤8h |  |  |  |  | Ref |
| $\leq 6$ | 0.11 | 0.04 | 2.62 | 0.008 | 1.12(1.03-1.22) |
| >8h | 0.08 | 0.07 | 1.19 | 0.234 | 1.09(0.95-1.25) |
| Diabetes |  |  |  |  |  |
| Diabetes=NO |  |  |  |  |  |
| 6~≤8h | | | | | Ref |
| $\leq 6$ | 0.17 | 0.04 | 4.70 | <.001 | 1.19(1.10-1.27) |
| >8h | 0.09 | 0.06 | 1.46 | 1.146 | 1.09(0.97-1.23) |
| Diabetes=Yes |  |  |  |  |  |
| 6~≤8h | | | | | Ref |
| $\leq 6$ | 0.21 | 0.07 | 3.07 | 0.002 | 1.24(1.08-1.42) |
| >8h | -0.09 | 0.13 | -0.64 | 0.519 | 0.91(0.71-1.19) |

Sensitivity analyses demonstrated that the main results remained consistent across various assessments. As illustrated in the Supplemental Table 1, the hazard ratio (HR) values were comparable to those obtained in the original analysis.

## Discussion

This study provides insights into the interplay between sleep duration, depression, and the risk of cardiovascular disease (CVD) in a Chinese national cohort. Our findings found that short sleep duration and depression were significantly associated with increased CVD risk. The associations remained significantly even after adjustment for other established cardiovascular risk factors. Our results also revealed a nonlinear relationship between sleep duration and cardiovascular events, as indicated by the Restricted Cubic Spline (RCS) analysis. This U-shaped association suggests that while short sleep duration is associated with increased CVD risk, longer sleep durations (beyond 10 hours) may also pose a risk. This finding aligns with existing literature, which indicates that both insufficient and excessive sleep can lead to adverse health outcomes[27]. Additionally, our mediation analysis highlighted that depression plays a significant role in the relationship between sleep duration and CVD risk. Combination of severe depression and short or long sleep duration further increased CVD risk. Our subgroup and sensitivity analyses further support the robustness of our findings.

There are several highlights in our study. Firstly, we utilized data from five waves of the CHARLS database, ensuring a sufficiently long follow-up period (2011-2020) for our analysis. Secondly, RCS plot was used to demonstrate the U-shape relationship between sleep duration and CVD risk. Thirdly, mediation analysis was performed to assess the role of depression in the relationship between sleep duration and CVD risk. Moreover, we also demonstrated the joint effect of sleep duration and depression on CVD risk. There is a fair amount of research reported that short sleep duration[28–30] and depression[8, 31] increased the risk of CVD. However, the interplay and combined effects of sleep duration and depression on CVD have been less studied. Combination effects of short or long sleep duration and depression on CVD risk was consistent with previous studies[27, 32]. However, sleep problems and depression are closely related. Our study extended this research by elucidating the mediated role of depression and the magnitude of its effect.

The mechanisms connecting sleep duration, depressive symptoms, and cardiovascular disease (CVD) remain complex and multifaceted, potentially involving both biological and behavioral pathways. Chronic inflammation may play a significant role; for instance, extended sleep duration has been shown to directly elevate low-grade inflammation[33]. Atypical depression is associated with various inflammatory cytokines, such as interleukin-6, C-reactive protein (CRP), and tumor necrosis factor (TNF)-α[34], all of which are critical contributors to atherosclerosis[35]. Conversely, short sleep duration is linked to hyperactivity of the hypothalamic-pituitary-adrenal (HPA) axis, evidenced by elevated cortisol levels. Dysregulation of the HPA axis is common in major depression[36], and hypercortisolemia has been associated with increased CVD risk and mortality[37].

While the potential shared genetic vulnerabilities affecting sleep, depression, and CVD have not been thoroughly explored, some studies suggest that genetic factors may partially explain the co-occurrence of these conditions[38]. Furthermore, obesity is a recognized risk factor for CVD that relates to both short and long sleep durations, as well as depression[39, 40]. Thus, the role of obesity in these interconnections warrants attention.

Additionally, both sleep duration and depression are linked to health-related behaviors that significantly influence CVD risk, such as smoking and physical inactivity[41–43]. For example, longer sleep duration is often associated with lower levels of physical activity[44], which negatively impacts body mass index and waist circumference, as well as exacerbating depressive symptoms. On the other hand, sleep restriction may diminish the likelihood of successful smoking cessation[45], a known risk factor for both depression and CVD.

More research is essential to elucidate the precise mechanisms by which sleep duration and depressive symptoms influence the development of CVD, as understanding these relationships could lead to targeted interventions for improving both mental and cardiovascular health.

Future research should aim to delve deeper into the mechanisms that mediate these associations. Longitudinal studies exploring the temporal relationships between sleep, depression, and CVD would provide valuable insights into causality. Additionally, it would be beneficial to investigate potential interventions, such as cognitive-behavioral therapy for insomnia (CBT-I) or sleep hygiene education, and their effectiveness in reducing both depressive symptoms and CVD risk.

Despite the robust findings, this study has limitations that must be acknowledged. The reliance on self-reported measures for sleep duration and depression may introduce bias, as individuals might misreport their sleep patterns or depression. However, previous studies have compared self-reported sleep durations with those measured by sleep diaries, actigraphy, and polysomnography. The findings indicated that self-reported sleep durations were well correlated with objective measurements[46, 47].While we attempted to control for various confounding factors, residual confounding cannot be entirely ruled out. Additionally, the generalizability of our findings may be limited to populations similar to those in our study, as cultural and lifestyle differences could influence sleep patterns and health outcomes.

In summary, our findings underscore the significant role of sleep duration in influencing cardiovascular health, particularly in the context of depression. Addressing both sleep and mental health is crucial for developing comprehensive strategies aimed at reducing the burden of cardiovascular diseases. Future studies should continue to explore these interactions and inform interventions that holistically address the multifaceted nature of health and well-being.

## Acknowledgements

The authors would like to express genuine gratitude to the participants and staff of the China Health and Retirement Longitudinal Study, who contributed greatly to the academic community and made this study possible.

## Availability of data and materials

The data that support the fndings of this study are available from the website of the China Health and Retirement Longitudinal Study (CHARLS) at http://charls.pku.edu.cn/.

## Declarations

### Ethics approval and consent to participate

CHARLS was ethically approved by the Institutional Review Board at Peking Uni versity (00001052-11014, 00001052-11015). All participants provided signed informed consent.

### Consent for publication

Not applicable.

## Reference

1. Hu SS, Writing Committee of the Report on Cardiovascular H, Diseases in C: Epidemiology and current management of cardiovascular disease in China. J Geriatr Cardiol 2024, 21(4):387–406.

2. Khot UN, Khot MB, Bajzer CT, Sapp SK, Ohman EM, Brener SJ, Ellis SG, Lincoff AM, Topol EJ: Prevalence of conventional risk factors in patients with coronary heart disease. JAMA 2003, 290(7):898–904.

3. Greenland P, Knoll MD, Stamler J, Neaton JD, Dyer AR, Garside DB, Wilson PW: Major risk factors as antecedents of fatal and nonfatal coronary heart disease events. JAMA 2003, 290(7):891–897.

4. Smith SC, Jr.: Current and future directions of cardiovascular risk prediction. Am J Cardiol 2006, 97(2A):28A–32A.

5. Global Cardiovascular Risk C, Magnussen C, Ojeda FM, Leong DP, Alegre-Diaz J, Amouyel P, Aviles-Santa L, De Bacquer D, Ballantyne CM, Bernabe-Ortiz A et al: Global Effect of Modifiable Risk Factors on Cardiovascular Disease and Mortality. N Engl J Med 2023, 389(14):1273–1285.

6. Ferrari AJ, Somerville AJ, Baxter AJ, Norman R, Patten SB, Vos T, Whiteford HA: Global variation in the prevalence and incidence of major depressive disorder: a systematic review of the epidemiological literature. Psychol Med 2013, 43(3):471–481.

7. Lei X, Sun X, Strauss J, Zhang P, Zhao Y: Depressive symptoms and SES among the mid-aged and elderly in China: evidence from the China Health and Retirement Longitudinal Study national baseline. Soc Sci Med 2014, 120:224–232.

8. Meng R, Yu C, Liu N, He M, Lv J, Guo Y, Bian Z, Yang L, Chen Y, Zhang X et al: Association of Depression With All-Cause and Cardiovascular Disease Mortality Among Adults in China. JAMA Netw Open 2020, 3(2):e1921043.

9. Li H, Zheng D, Li Z, Wu Z, Feng W, Cao X, Wang J, Gao Q, Li X, Wang W et al: Association of Depressive Symptoms With Incident Cardiovascular Diseases in Middle-Aged and Older Chinese Adults. JAMA Netw Open 2019, 2(12):e1916591.

10. Cui Y, Zhu C, Lian Z, Han X, Xiang Q, Liu Z, Zhou Y: Prospective association between depressive symptoms and stroke risk among middle-aged and older Chinese. BMC Psychiatry 2021, 21(1):532.

11. Huang T, Mariani S, Redline S: Sleep Irregularity and Risk of Cardiovascular Events: The Multi-Ethnic Study of Atherosclerosis. J Am Coll Cardiol 2020, 75(9):991–999.

12. Yan B, Yang J, Zhao B, Fan Y, Wang W, Ma X: Objective Sleep Efficiency Predicts Cardiovascular Disease in a Community Population: The Sleep Heart Health Study. J Am Heart Assoc 2021, 10(7):e016201.

13. Hsieh CG, Martin JL: Short Sleep, Insomnia, and Cardiovascular Disease. Curr Sleep Med Rep 2019, 5(4):234–242.

14. Cappuccio FP, Cooper D, D’Elia L, Strazzullo P, Miller MA: Sleep duration predicts cardiovascular outcomes: a systematic review and meta-analysis of prospective studies. Eur Heart J 2011, 32(12):1484–1492.

15. Javaheri S, Redline S: Insomnia and Risk of Cardiovascular Disease. Chest 2017, 152(2):435–444.

16. Somers VK, White DP, Amin R, Abraham WT, Costa F, Culebras A, Daniels S, Floras JS, Hunt CE, Olson LJ et al: Sleep apnea and cardiovascular disease: an American Heart Association/American College of Cardiology Foundation Scientific Statement from the American Heart Association Council for High Blood Pressure Research Professional Education Committee, Council on Clinical Cardiology, Stroke Council, and Council on Cardiovascular Nursing. J Am Coll Cardiol 2008, 52(8):686–717.

17. Liu C, Ye Z, Chen L, Wang H, Wu B, Li D, Pan S, Qiu W, Ye H: Interaction effects between sleep-related disorders and depression on hypertension among adults: a cross-sectional study. BMC Psychiatry 2024, 24(1):482.

18. Zhang D, Qu Y, Zhai S, Li T, Xie Y, Tao S, Zou L, Tao F, Wu X: Association between healthy sleep patterns and depressive trajectories among college students: a prospective cohort study. BMC Psychiatry 2023, 23(1):182.

19. Baglioni C, Battagliese G, Feige B, Spiegelhalder K, Nissen C, Voderholzer U, Lombardo C, Riemann D: Insomnia as a predictor of depression: a meta-analytic evaluation of longitudinal epidemiological studies. J Affect Disord 2011, 135(1-3):10–19.

20. Zhao Y, Hu Y, Smith JP, Strauss J, Yang G: Cohort profile: the China Health and Retirement Longitudinal Study (CHARLS). Int J Epidemiol 2014, 43(1):61–68.

21. Itani O, Jike M, Watanabe N, Kaneita Y: Short sleep duration and health outcomes: a systematic review, meta-analysis, and meta-regression. Sleep Med 2017, 32:246–256.

22. Liu Y, Wang C, Liu Y: Association between adverse childhood experiences and later-life cardiovascular diseases among middle-aged and older Chinese adults: The mediation effect of depressive symptoms. J Affect Disord 2022, 319:277–285.

23. Liu Y, Ning N, Sun T, Guan H, Liu Z, Yang W, Ma Y: Association between solid fuel use and nonfatal cardiovascular disease among middle-aged and older adults: Findings from The China Health and Retirement Longitudinal Study (CHARLS). Sci Total Environ 2023, 856(Pt 2):159035.

24. Cao Z, Hou Y, Yang H, Huang X, Wang X, Xu C: Healthy sleep patterns and common mental disorders among individuals with cardiovascular disease: A prospective cohort study. J Affect Disord 2023, 338:487–494.

25. Song J, Chen X, Jiang Y, Mi J, Zhang Y, Zhao Y, Wu X, Gao H: Association and Interaction Analysis of Lipid Accumulation Product with Impaired Fasting Glucose Risk: A Cross-Sectional Survey. J Diabetes Res 2019, 2019:9014698.

26. Cui C, Liu L, Zhang T, Fang L, Mo Z, Qi Y, Zheng J, Wang Z, Xu H, Yan H et al: Triglyceride-glucose index, renal function and cardiovascular disease: a national cohort study. Cardiovasc Diabetol 2023, 22(1):325.

27. Zhu C, Wang J, Wang J, Zhong Q, Huang Y, Chen Y, Lian Z: Associations between depressive symptoms and sleep duration for predicting cardiovascular disease onset: A prospective cohort study. J Affect Disord 2022, 303:1–9.

28. Han H, Wang Y, Li T, Feng C, Kaliszewski C, Su Y, Wu Y, Zhou J, Wang L, Zong G: Sleep Duration and Risks of Incident Cardiovascular Disease and Mortality Among People With Type 2 Diabetes. Diabetes Care 2023, 46(1):101–110.

29. Kwok CS, Kontopantelis E, Kuligowski G, Gray M, Muhyaldeen A, Gale CP, Peat GM, Cleator J, Chew-Graham C, Loke YK et al: Self-Reported Sleep Duration and Quality and Cardiovascular Disease and Mortality: A Dose-Response Meta-Analysis. J Am Heart Assoc 2018, 7(15):e008552.

30. Daghlas I, Dashti HS, Lane J, Aragam KG, Rutter MK, Saxena R, Vetter C: Sleep Duration and Myocardial Infarction. J Am Coll Cardiol 2019, 74(10):1304–1314.

31. Krittanawong C, Maitra NS, Qadeer YK, Wang Z, Fogg S, Storch EA, Celano CM, Huffman JC, Jha M, Charney DS et al: Association of Depression and Cardiovascular Disease. Am J Med 2023, 136(9):881–895.

32. Deschenes SS, Burns RJ, Graham E, Schmitz N: Depressive symptoms and sleep problems as risk factors for heart disease: a prospective community study. Epidemiol Psychiatr Sci 2019, 29:e50.

33. Irwin MR, Olmstead R, Carroll JE: Sleep Disturbance, Sleep Duration, and Inflammation: A Systematic Review and Meta-Analysis of Cohort Studies and Experimental Sleep Deprivation. Biol Psychiatry 2016, 80(1):40–52.

34. Milaneschi Y, Lamers F, Berk M, Penninx B: Depression Heterogeneity and Its Biological Underpinnings: Toward Immunometabolic Depression. Biol Psychiatry 2020, 88(5):369–380.

35. Lutgens E, Atzler D, Doring Y, Duchene J, Steffens S, Weber C: Immunotherapy for cardiovascular disease. Eur Heart J 2019, 40(48):3937–3946.

36. Lamers F, Vogelzangs N, Merikangas KR, de Jonge P, Beekman AT, Penninx BW: Evidence for a differential role of HPA-axis function, inflammation and metabolic syndrome in melancholic versus atypical depression. Mol Psychiatry 2013, 18(6):692–699.

37. Juruena MF, Bocharova M, Agustini B, Young AH: Atypical depression and non-atypical depression: Is HPA axis function a biomarker? A systematic review. J Affect Disord 2018, 233:45–67.

38. Bondy B: Common genetic factors for depression and cardiovascular disease. Dialogues Clin Neurosci 2007, 9(1):19–28.

39. Ogilvie RP, Patel SR: The epidemiology of sleep and obesity. Sleep Health 2017, 3(5):383–388.

40. Patist CM, Stapelberg NJC, Du Toit EF, Headrick JP: The brain-adipocyte-gut network: Linking obesity and depression subtypes. Cogn Affect Behav Neurosci 2018, 18(6):1121–1144.

41. Weinberger AH, Kashan RS, Shpigel DM, Esan H, Taha F, Lee CJ, Funk AP, Goodwin RD: Depression and cigarette smoking behavior: A critical review of population-based studies. Am J Drug Alcohol Abuse 2017, 43(4):416–431.

42. Purani H, Friedrichsen S, Allen AM: Sleep quality in cigarette smokers: Associations with smoking-related outcomes and exercise. Addict Behav 2019, 90:71–76.

43. Stefan L, Vrgoc G, Rupcic T, Sporis G, Sekulic D: Sleep Duration and Sleep Quality Are Associated with Physical Activity in Elderly People Living in Nursing Homes. Int J Environ Res Public Health 2018, 15(11).

44. Stranges S, Dorn JM, Shipley MJ, Kandala NB, Trevisan M, Miller MA, Donahue RP, Hovey KM, Ferrie JE, Marmot MG et al: Correlates of short and long sleep duration: a cross-cultural comparison between the United Kingdom and the United States: the Whitehall II Study and the Western New York Health Study. Am J Epidemiol 2008, 168(12):1353–1364.

45. Rapp K, Buechele G, Weiland SK: Sleep duration and smoking cessation in student nurses. Addict Behav 2007, 32(7):1505–1510.

46. Lauderdale DS, Knutson KL, Yan LL, Liu K, Rathouz PJ: Self-reported and measured sleep duration: how similar are they? Epidemiology 2008, 19(6):838–845.

47. Silva GE, Goodwin JL, Sherrill DL, Arnold JL, Bootzin RR, Smith T, Walsleben JA, Baldwin CM, Quan SF: Relationship between reported and measured sleep times: the sleep heart health study (SHHS). J Clin Sleep Med 2007, 3(6):622–630.

